# Life-course neighbourhood socioeconomic disadvantage and atherosclerotic carotid artery plaques. The Cardiovascular Risk in Young Finns Study

**DOI:** 10.1101/2025.07.07.25331069

**Authors:** Olli Raitakari, Jaana Pentti, Juhani S. Koskinen, Juha Mykkänen, Suvi Rovio, Katja Pahkala, Markus Juonala, Terho Lehtimäki, Mika Kähönen, Ichiro Kawachi, Mika Kivimäki, Jorma Viikari, Jussi Vahtera

## Abstract

**Background:** Neighbourhood socioeconomic disadvantage is a known determinant of cardiovascular disease (CVD) risk. However, its impact on subclinical atherosclerosis across the life course remains inadequately understood. This study examined the association between cumulative neighbourhood socioeconomic disadvantage from childhood to midlife and carotid artery plaques— a marker of subclinical atherosclerosis—independent of genetic and behavioral CVD risk factors.

**Methods:** We analysed data from 2,051 participants in the Cardiovascular Risk in Young Finns Study, a prospective cohort followed from childhood (mean age 10.7 years in 1980) to adulthood (mean age 48.6 years in 2018–2020). Neighbourhood disadvantage was derived from national grid-based socioeconomic data and computed cumulatively across childhood/adolescence, adulthood, and the entire life course. The number of carotid artery plaques (plaque count) were assessed by standardized ultrasound imaging. Multivariable Poisson regression models were used to evaluate associations, adjusting for age, sex, individual and parental socioeconomic status, genetic predisposition, and cardiovascular risk profiles. Mediation analyses assessed the role of ideal cardiovascular health (CVH) metrics.

**Results:** No cross-sectional association was found between current neighbourhood disadvantage and carotid plaque count. However, higher cumulative neighbourhood disadvantage over the life course was associated with increased plaque count (rate ratio [RR] ≈ 1.20 per 1 SD increase). This relationship persisted after controlling for parental carotid artery plaques, polygenic coronary artery disease risk score, and Framingham risk score. The effect was partially mediated by ideal CVH metrics, particularly smoking and blood pressure, which collectively explained up to 50% of the association.

**Conclusions:** Long-term exposure to neighbourhood socioeconomic disadvantage beginning in childhood is associated with subclinical atherosclerosis in midlife independently of achieved socioeconomic position. Behavioural risk factors partially mediate this link, highlighting the importance of early and sustained interventions targeting both social environments and health behaviours to mitigate cardiovascular risk.

## Introduction

Neighbourhood socioeconomic disadvantage has been shown to be associated with higher risk of cardiovascular diseases, such as coronary artery disease and stroke^1,2^. In the Atherosclerosis Risk in Communities Study, for example, individuals living in disadvantaged neighbourhoods had increased incidence of coronary artery disease^1^. Similarly, in the Multi-Ethnic Study of Atherosclerosis, social disadvantage, measured using a weighted aggregate score including neighbourhood and physical environment, was associated with a 31% higher risk of total cardiovascular disease over median follow-up of 14 years ^3^. In most cases, these cardiovascular disease outcomes are manifestations of the end stages of atherosclerosis affecting the walls of medium-sized and large-sized arteries - a disease that often begins early in life and progresses asymptomatically for decades^4,5^.

Since atherosclerosis develops gradually over the life course, cohort studies focusing on atherosclerosis can provide valuable insights into the long-term effects of neighbourhood context. Understanding the link between neighbourhood socioeconomic disadvantage, beginning from childhood, and early markers of atherosclerosis in adulthood can provide insights into the origins and progression of the disease. Ultrasound imaging of carotid arteries provides a non-invasive method to measure atherosclerotic plaques in epidemiologic studies. Carotid artery plaques are markers of generalized atherosclerosis and sources of thromboemboli^6^, and they are associated with approximately twofold increased risk of coronary artery disease and stroke^7–9^. Studying asymptomatic plaques allows for the identification of individuals at higher risk of developing clinical cardiovascular outcomes. This can open a window for earlier intervention and prevention strategies^4,10^. In addition, investigating the association at the subclinical stage can help elucidate the pathways through which neighbourhood socioeconomic disadvantage over the life course influences cardiovascular health. Furthermore, while individual-level socioeconomic status is a strong predictor of cardiovascular outcomes, studying subclinical markers helps determine if neighbourhood socioeconomic disadvantage has an independent effect on atherosclerosis after accounting for individual-level socioeconomic factors. The independent role of neighbourhood disadvantage in increasing cardiovascular risk is still uncertain, as many studies have observed that the association between neighbourhood socioeconomic status and cardiovascular disease risk attenuates or loses statistical significance after accounting for individual socioeconomic status^11–13^. Moreover, most studies have examined neighbourhood environments at a single point in the life course. Only a few studies have examined neighbourhood exposures across the life course, and therefore more research is needed to understand how neighbourhood environments during different stages of life affect cardiovascular health^14^. Thus, while the association between neighbourhood socioeconomic disadvantage and clinical cardiovascular disease is established, studying asymptomatic carotid artery plaques offers a critical lens for understanding the early development of atherosclerosis, identifying at-risk populations for primary prevention, and elucidating the specific pathways through which neighbourhood disadvantage may impact cardiovascular health across the life course. Understanding these complex relationships is essential for developing targeted, effective interventions to reduce cardiovascular health disparities associated with neighbourhood environments.

## Methods

The Cardiovascular Risk in Young Finns Study (YFS) is a prospective multicentre study from Finland initiated in the late 70’s. The first large baseline examination was conducted in 1980 (baseline age, 3–18 years, N=3,596)^15^. Children aged 3, 6, 9, 12, 15, and 18 years were randomly chosen from the population register from the five Finnish university cities with medical faculties and their surrounding rural communities. The cohort has been followed up with large-scale field studies in 1983, 1986, 1989, 1992, 2001, 2007, 2011, and 2018-20. The participation rates have varied between 61-85% depending on the data collection. The latest follow-up was extended to cover the original participants as well as their parents and offspring aged 3 years and older. This multigenerational follow-up of the cohort was executed from March 2018 to February 2020. We contacted the remaining, *i.e.* those alive and living in Finland, original participants (N=3,309) to inform them about the beginning of the new multigenerational data collection. At this time, they had the possibility to decline the offer to invite their parents and/or offspring to participate in the new study. After excluding those who were deceased, decided to withdraw from the study, and participants with missing contact information, we invited 3,217 original participants (Generation 1; G1, aged 40-58 y). Additionally, we invited 5,696 offspring (Generation 2; G2, 3-38 y), and 3,940 parents (Generation 0; G0, 59-92 y) of the original participants, including parents and adult offspring of those original participants who were deceased. In total, the number of invited individuals was 12,853. Altogether 7,341 of the invited individuals provided any data (57.1%). A total of 6,753 (52.5%) participants attended the clinical examination, while 588 participants provided questionnaire data only. The generation-specific participation rates for participants with any data were 66.1% for G1 (N=2,127; females 55.0%), 62.2% for G0 (N=2,452; females 61.1%), and 48.5% for G2 (N=2,762; females 54.9%). The details of the study design and methods have been published^16^. In the present analyses, we used data on the original cohort members (G1) and their parents (G0).

### Clinical examination and questionnaires

Height and weight were measured at the clinic visit, and body mass index (BMI; kg/m2) was calculated. Blood pressure was measured three times in a sitting position from the right arm brachial artery by an automatic Omron HBP-1300 blood pressure measurement device (Omron Corporation, Kyoto, Japan). The mean of these measurements was calculated. Data on medical conditions diagnosed by a physician, medications, smoking habits and alcohol consumption were collected by self-report questionnaires. Information on clinical cardiovascular diagnoses was collected using a self-report questionnaire and also by using national health database for G1.

### Biochemical measurements

Venous blood samples for biomarker analyses were obtained after a minimum 4 hour fast and stored at -70°C until analysis. Serum total cholesterol, HDL-cholesterol, and triglycerides were determined with enzymatic assays on an Indiko Plus analyzer (Thermo Fisher Scientific, Finland). LDL-cholesterol was estimated with the Friedewald’s formula when level of triglycerides was <4.0 mmol/l. Non-HDL-cholesterol was calculated as total cholesterol - HDL-cholesterol.

### Definition of type 2 diabetes and hypertension

Participants were classified as having type 2 diabetes if at least one of the following criteria was fulfilled 1) fasting glucose ≥ 7.0 mmol/l, 2) HbA1c ≥ 6.5% (47.55 mmol/mol), 3) self-reported diagnosis of type 2 diabetes, 4) self-reported use of diabetes medication, or 5) use of medication for type 2 diabetes based on the register data of the Social Insurance Institution of Finland. Participants were classified as having hypertension if at least one of the following criteria was fulfilled 1) systolic blood pressure ≥140 mmHg, 2) diastolic blood pressure ≥90, 3) self-reported diagnoses for hypertension, 4) self-reported medication of hypertension.

### Carotid ultrasonography

Carotid ultrasound studies were performed using General Electric (GE) Logiq S8 (GE Vingmed Ultrasound A/S, Horten, Norway) ultrasound mainframes with a ML6-15-D matrix linear transducer. Carotid ultrasound studies were performed according to standardized protocols by sonographers and trained ultrasound technicians to assess left and right carotid artery plaques and intima-media thickness (IMT) of the common carotid artery, carotid bifurcation, and internal carotid artery. Participants were examined in supine position with their necks extended. During the ultrasonography, a continuous electrocardiogram was recorded. The detailed methods have been published^16^.

### Measurement of carotid plaques

During ultrasonography, left and right carotid arteries were scanned for carotid artery plaques, defined as a focal structure protruding into the arterial lumen of at least 0.5 mm or 50% of the surrounding IMT value or demonstrating a thickness of at least 1.5 mm as measured from the media-adventitia interface to the intima-lumen interface^17^.

When a carotid plaque was suspected, it was scanned from the longitudinal view to ensure accurate visualization of the plaque-adventitia and plaque-lumen interfaces. A 5-second moving clip was then captured and stored for subsequent off-line plaque analysis. For all participants, moving cross-sectional scans of both the left and right carotid arteries were taken, starting from the common carotid artery, and extending to the point where the internal and external carotid arteries branched. These scans were stored for off-line plaque analysis. Plaques were recorded in the common carotid artery, carotid bifurcation, and internal carotid artery. The total plaque count was calculated as the sum of all plaques for each carotid segment, derived from the longitudinal view.

### Neighbourhood socioeconomic disadvantage

Data regarding neighbourhood social disadvantage was derived from a grid database established and maintained by Statistics Finland. The database contains socio-economic information from each residence at a spatial resolution of 250 meters by 250 meters^18^. The grid data were obtained for the years 1990 (the first year these data were available from Statistics Finland), 1995 and at 2-year intervals from 2000 to 2020. The neighbourhood disadvantage score is based on the proportion of adults with low education, the unemployment rate, and the average annual income of households in each 250m x 250m grid area. Missing data (*i.e.* areas with fewer than 10 residents in the neighbourhood) were replaced with the mean neighbourhood disadvantage score of the eight adjacent map squares. For each of the three variables, we derived a standardized z score based on the total Finnish population (mean=0, SD=1). A score for neighbourhood disadvantage was then calculated by taking the mean value across the three z scores. Higher scores on the continuous index denote greater disadvantage. For the statistical analyses, the continuous neighbourhood disadvantage score was also classified into two categories using the national means as the cut-off point.

High quality residential mobility data, based on a complete history of the residential addresses with latitude and longitude coordinates between years 1980 (the baseline) and 2018 (the last follow-up), were obtained from the Population Register Centre for each participant. Data on the residential neighbourhood disadvantage for each time point were linked to the cohort participants’ home addresses using latitude and longitude coordinates. A cumulative socioeconomic disadvantage score weighted by residential time at each location in between 1980 and 2018 was calculated for each participant.

### Cardiovascular risk factors

#### Parental plaque load

Carotid plaques were measured from the parents of 1,025 participants and included data from 590 fathers and 896 mothers. The mean age of fathers was 73.4 years (SD 5.4), and their mean number of carotid plaques (plaque count) was 3.39 (SD 2.45). The mean age of mothers was 72.4 years (SD 5.9), and their mean plaque count was 2.20 (SD 1.92). The parents were divided into two groups using age 72 years as a cut-off point. Participants were classified as having a high parental plaque load if either parent had carotid plaques above the median for their age and sex-specific age-group (<72 years: >2 plaques in mothers and fathers; >72 years: >4 plaques in mothers and >5 plaques in fathers).

#### Polygenic risk score for coronary artery disease

Genotyping was performed using either a custom build Illumina Human 670K BeadChip (670K) for 2,443 samples collected in 2001, 2007, and 2011 follow-up studies or with Illumina Infinium Global Screening Array (GSA) for 403 samples collected in 2018-2020 follow-up study. The 670K genotypes were called using Illuminus clustering algorithm^19^, whereas the GSA genotypes were called using Illumina’s GenCall algorithm. The following filters were used for sample and SNP quality control for all genotype data: sample and SNP call rate <0·95, cryptic relatedness (pi-hat >0·2), SNP Hardy-Weinberg equilibrium test (p ≤ 1×10-6). Samples with sex discrepancy, excess heterozygosity, as well as genetic outliers detected with multidimensional scaling, were excluded. Genotype imputation was performed using Minimac3 and Trans-Omics in Precision Medicine Initiative (TOPMed) r3 reference set on the TOPMed Imputation Server. Polygenic risk score for coronary artery disease (CAD PRS ) was calculated with PRS-CS software that infers posterior SNP effect sizes under continuous shrinkage priors using GWAS summary statistics and an external LD reference panel^20^. The results from UK Biobank and CARDIoGRAMplusC4D meta-analysis were used as CAD GWAS summary statistics^21^, and UK Biobank European data was used as a LD reference. PRS-CS software was run with default options. The posterior SNP effect sizes (weights) from PRS-CS were then harmonized to include the same variants (N=603,448) across G1 and G0. Finally, the CAD PRS was calculated with ‘--score’-function of PLINK2 software and scaled to standard deviation (SD) unit. For statistical analyses the CAD PRS was dichotomised to low (below median) to high (at or above median).

#### Framingham risk score

The Framingham risk score in 2018 was used to identify participants at high risk of developing coronary artery disease (score ≥7.5%). The algorithm estimates 10-year coronary artery disease risk separately for men and women on the basis of age, total cholesterol, HDL-cholesterol, systolic blood pressure, and cigarette smoking^22^.

#### Ideal cardiovascular health

The ideal cardiovascular health score was determined based on the American Heart Association ideal cardiovascular health metrics including ideal health behaviours (BMI <25 kg/m2, non-smoking, ideal diet based on recommendations, and physical activity at goal level) as well as ideal health factors (systolic blood pressure <120 mmHg, total cholesterol level <5.17, and fasting plasma glucose <5.55)^23^. Individuals with summed score of three or higher were defined as having ideal or intermediate cardiovascular health score.

### Covariates

In addition to age and sex, the participants’ own socioeconomic characteristics used in the study included education (basic or vocational vs. higher), marital status (married or cohabiting vs. other), childhood socioeconomic position (which was defined by parental socioeconomic position during their childhood), number of moves between 1980 and 2018, and density between 1980 and 2018 (mean number of residents in the neighbourhoods). Parental socioeconomic status (SES) was defined on the grounds of the level of parental education, mean household income and unemployment (yes or no). Standardized scores (mean 0, SD 1) of aforementioned education and income variables, and unemployment score (-1 for history of unemployment and 0 for no unemployment) were summed up and the mean score ≤0 / >0 was used to define high versus low parental SES. Information on density was derived from the Statistics Finland grid database.

### Statistical methods

We determined the participants’ exposure to cumulative neighbourhood socioeconomic disadvantage between 1980 and 2018, separately for childhood/adolescence (ages 6 to 20 years), adulthood (ages 21 to 55) and the whole life course (ages 6 to 55) by summing up the residential time-weighted disadvantage scores over time during each life stage. Current disadvantage was based on neighbourhood socioeconomic disadvantage in the Jan 1, 2018 residential location. In the analyses on the number of carotid plaques (plaque count), we used Poisson regression analyses and expressed the results as rates (average number of plaques per individual), and rate ratios and their 95% confidence intervals (95% CI).

First, we examined the associations between cumulative neighbourhood socioeconomic disadvantage (treated as a continuous variable) during the three different life stages, as well as current residential neighbourhood and the number of carotid plaques. The models were adjusted for age and sex and additionally for parental SES (for childhood/adolescence exposure) or parental SES, own education and marital status (in analyses of adulthood and whole life course exposure). Second, we examined the associations between CVD risk factors (high or low), including parental plaque load, CAD PRS, Framingham risk score and ideal CVH score – and the number of carotid plaques adjusted for age and sex. CAD PRS risk score was also adjusted for genetic principal components. Additionally, these models were adjusted for parental SES, own education, marital status, number of moves, and population density in the neighbourhood.

In the main analyses, we examined the associations of high vs low cumulative neighbourhood disadvantage (standardized z score based on the total Finnish population >0 or ≤0) between 1980 and 2020 among participants based on their CVD risk status. In these analyses, participants were categorized into groups: (1) low/high neighbourhood disadvantage and low/high parental plaque load, (2) low/high neighbourhood disadvantage and low/high CAD PRS, (3) low/high neighbourhood disadvantage and low/high Framingham risk score and (4) low/high neighbourhood disadvantage and ideal/intermediate CVH score. From these analyses, we report the plaque counts (95% CIs) for each group as well as the rate ratio (95% Cis) of high vs. low neighbourhood disadvantage in the statuses of each low and high CVD within each stratum. As earlier, the models were adjusted for age and sex (and genetic principal components for CAD PRS) and additionally, for parental SES, own education and marital status.

Finally, to identify the risk indicators (ideal CVH score and its components) that mediate the associations between neighbourhood disadvantage and carotid plaques, we estimated the proportion of this association mediated by the ideal CVH score components in combination, and each component separately, including blood pressure, cholesterol, glucose, smoking, physical activity, BMI and diet. Confidence intervals were estimated by bootstrapping with 500 samples. Specifically, we used an inverse odds ratio-weighted method to estimate the extent to which the exposure (here neighbourhood disadvantage) directly affects the outcome when excluding the mediator pathway^24^. The inverse odds ratio-weighted method allows simultaneous assessment of multiple mediators, as well as individual mediators, by regressing the set of mediators on the exposure. The causal effects were divided into direct effects, indirect effects, and total effects.

## Results

Of the 3,596 baseline participants in 1980, 1,534 (43%) did not participate in the carotid ultrasound examination in 2018-20. Compared to those who participated, they were slightly younger at baseline (10.1 vs 10.7 years), more often boys (54% vs 45%), had a low parental SES (60% vs. 52%) and their mean neighbourhood disadvantage in childhood and adolescence was higher (0.11 vs. -0.03).

Of the 2,062 cohort members, who participated in the 2018-20 ultrasound examination, 1,998 (97%) provided data on their cumulative neighbourhood socioeconomic disadvantage between 1980 and 2020. The mean age was 10.7 years (range 3-18) at baseline in 1980 and 48.6 years (range 41-56) at the end of the 38-year follow-up in 2018-2020. Mean number (*i.e.* count) of carotid plaques by the characteristics of the study population are shown in Table 1. Higher rates were observed in relation to male sex, low education, low parental SES, high parental plaque load, high CAD PRS, high Framingham risk score, poor ideal CVH total score and all non-ideal CVH score components. The mean residential time across the life course was 38.3 years). Participants classified into high and low neighbourhood disadvantage categories (based on their cumulative score from 1980 to 2018 using the national mean as the cut-off point) did not differ in terms of age, sex, CAD PRS, or parental carotid plaque load.

**Table 1.**
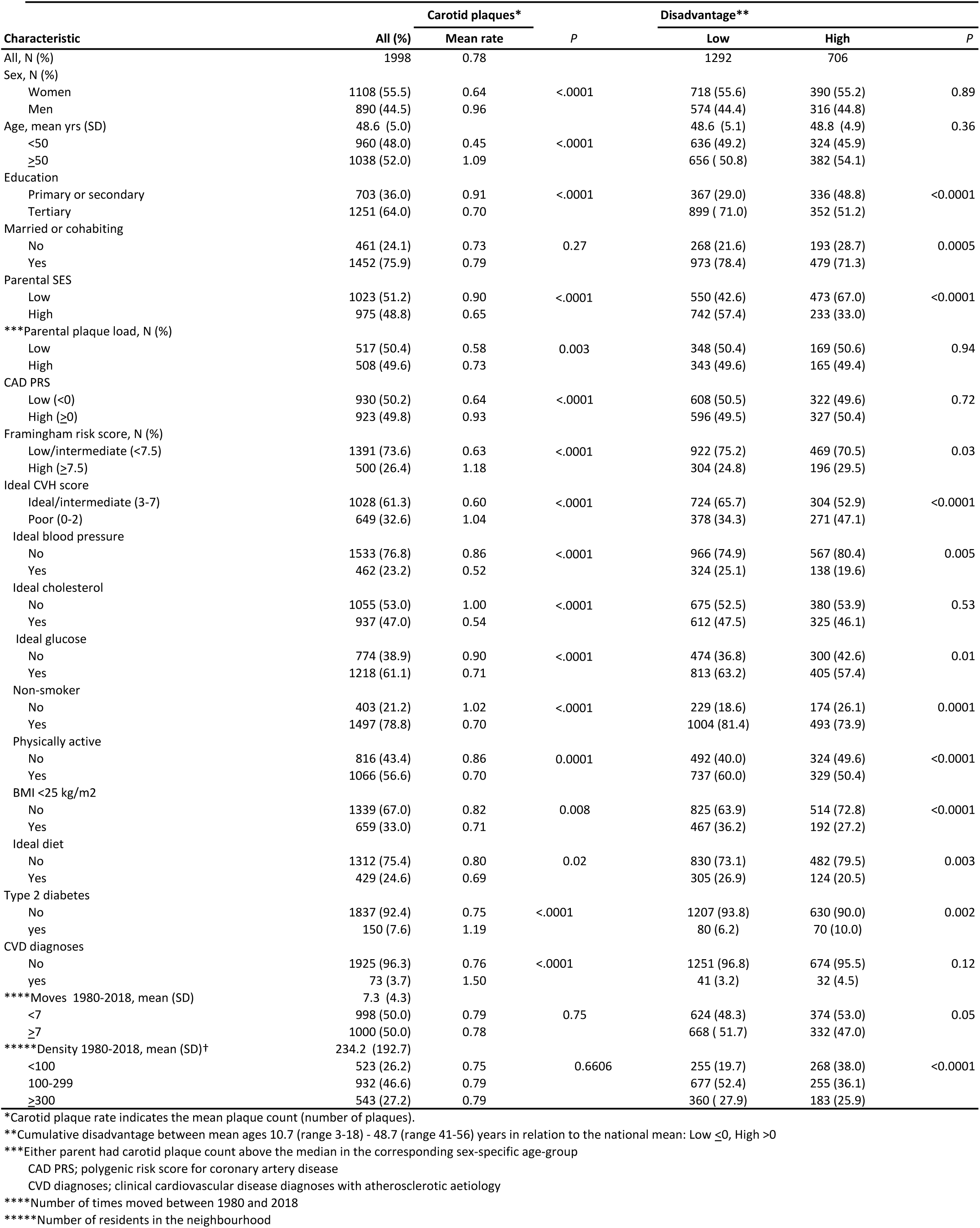
Characteristics of the participants.

Individuals exposed to high cumulative neighbourhood socioeconomic disadvantage had on average lower own education level and lower parental socioeconomic position compared to individuals exposed to low disadvantage. They were also less often married or cohabiting, had higher prevalence of type 2 diabetes (10.0 vs. 6.2%, p=0.002) and atherosclerotic cardiovascular diseases (4.5 vs. 3.2%, p=0.12), and (Table 1).

High neighbourhood socioeconomic disadvantage was also associated with higher cardiovascular risk, as assessed by the Framingham risk score, and inversely associated with the ideal CVH score. The distributions of all individual components of the ideal CVH score differed between the two groups, except for ideal cholesterol level (Table 1).

The associations between cumulative neighbourhood socioeconomic disadvantage across life-stages and the midlife carotid plaques are shown in Table 2. Four distinct exposure periods were examined: 1) cumulative socioeconomic disadvantage over the life course; 2) cumulative neighbourhood socioeconomic disadvantage during childhood/adolescence; 3) cumulative neighbourhood socioeconomic disadvantage during adulthood; and 4) current neighbourhood socioeconomic disadvantage, *i.e.* at the time of the carotid plaques assessment. No cross-sectional associations were observed between the current neighbourhood disadvantage and plaques.

**Table 2.**
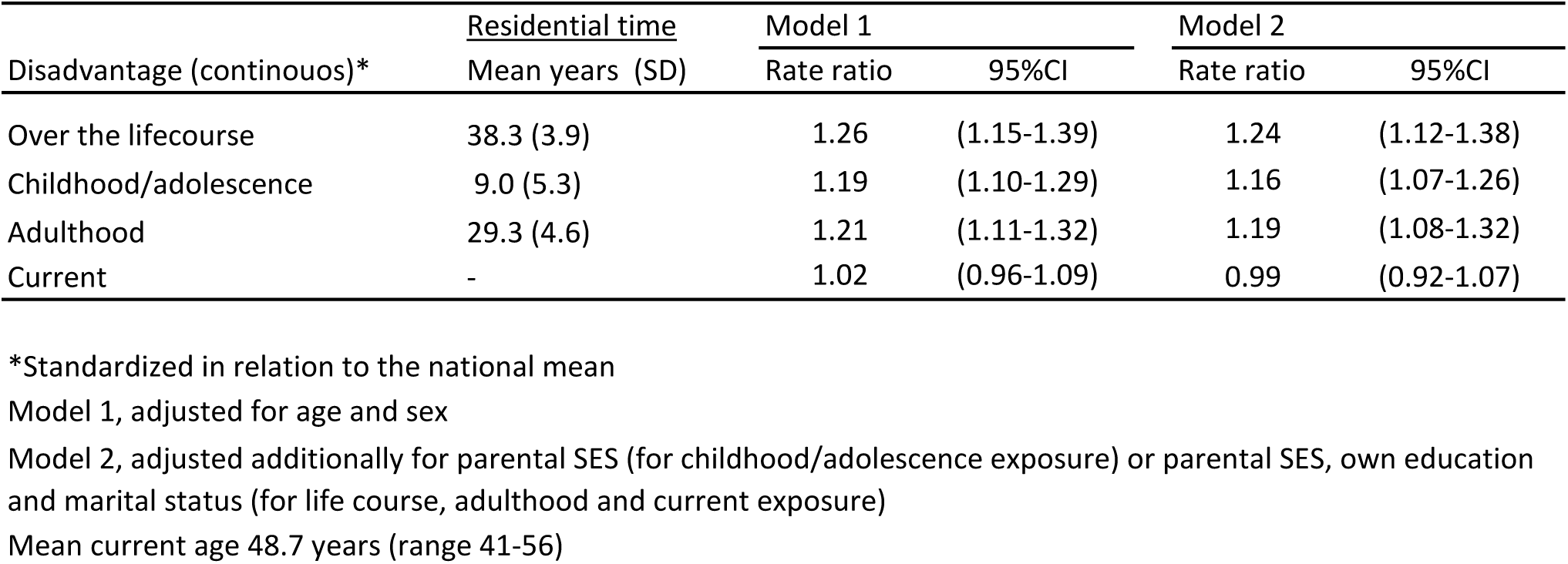
Cumulative neighbourhood disadvantage at different life stages and the number of carotid plaques (rate ratio / 1 SD increase in disadvantage)

However, cumulative disadvantage exposure was significantly associated with higher carotid plaque count in models adjusted for age and sex, and additionally for own and/or parental socioeconomic status, own marital status, as well as the number of moves and population density between 1980 and 2018. The strongest associations between neighbourhood disadvantage and plaques were observed in the whole life course models. The rate ratios indicated about 20% increase in plaque count by every 1 standard deviation increase in cumulative disadvantage.

Both genetic risk markers (parental plaque load and CAD PRS) and lifestyle risk markersa (Framingham score and ideal CVH score) were directly associated with carotid plaque count (Table 3). For example, above the median values for CAD PRS was associated with about 35% higher plaque count as compared to below median values. A high Framingham risk score (over 7.5%) was associated with approximately 45% higher carotid plaque count compared low to a low Framingham risk score. Similarly, having poor ideal CVH score, as opposed to an ideal or intermediate score was also associated with an approximately 45% higher plaque count.

**Table 3.** Associations of CVD risk factors and cumulative neighbourhood disadvantage with carotid plaques.

| Risk factor | Adjustement | N | Plaque |  | RR | (95% CI) |
| --- | --- | --- | --- | --- | --- | --- |
|  |  |  | count | (95% CI) |  |  |
| Parental plaque load | Age and sex |  |  |  |  |  |
| Low |  | 517 | 0.48 | (0.43-0.54) | 1.00 |  |
| High |  | 508 | 0.67 | (0.60-0.74) | 1.38 | (1.19-1.61) |
| CAD PRS | Age, sex, genetic principal components |  |  |  |  |  |
| Low (<0 SD) |  | 930 | 0.63 | (0.57-0.70) | 1.00 |  |
| High (≥0) |  | 923 | 0.86 | (0.77-0.95) | 1.36 | (1.22-1.51) |
| Framingham risk score | Age and sex |  |  |  |  |  |
| Low (<7.5) |  | 1391 | 0.61 | (0.57-0.65) | 1.00 |  |
| High (≥7.5) |  | 500 | 0.88 | (0.80-0.97) | 1.45 | (1.28-1.64) |
| Ideal CVH score | Age and sex |  |  |  |  |  |
| Ideal or intermediate (3-7) |  | 1028 | 0.57 | (0.52-0.62) | 1.00 |  |
| Poor (0-2) |  | 649 | 0.83 | (0.76-0.90) | 1.45 | (1.30-1.63) |
| Neighborhood disadvantage* | Age and sex |  |  |  |  |  |
| Low (≤0) |  | 1292 | 0.63 | (0.59-0.68) | 1.00 |  |
| High (>0) |  | 706 | 0.80 | (0.73-0.87) | 1.26 | (1.14-1.40) |
| High | +Parental plaque load |  |  |  | 1.20 | (1.02-1.40) |
| High | +Polygenic CAD risk score |  |  |  | 1.25 | (1.12-1.38) |
| High | +Framingham risk score |  |  |  | 1.24 | (1.11-1.38) |
| High | +Ideal CVH score |  |  |  | 1.15 | (1.02-1.29) |

\*Cumulative disadvantage between mean ages 10.7-48.7 years in relation to the national mean: Low <0, High >0 CAD PRS; polygenic risk score for coronary artery disease

The association between high vs. low cumulative neighbourhood disadvantage and carotid plaques (RR=1.26) remained statistically significant and was not substantially attenuated in models additionally adjusted for parental plaque load, CAD PRS and Framingham risk score (RR after adjustments 1.24-1.25). The association was similar in both sexes: men RR=1.25 (1.11-1.42), women 1.27 (1.10-1.47), sex-interaction p=0.89. When the ideal CVH score was included in the model, the association between neighbourhood disadvantage and plaque remained significant, but was attenuated to RR=1.15. This suggests that the effect of neighbourhood disadvantage on carotid plaques may be partially mediated through cardiovascular health captured by the ideal CVH score.

To further examine how the background risk factor profile may influence the link between neighbourhood disadvantage and carotid plaques, we examined this association for the combinations of low/high neighbourhood disadvantage and low/high background risk (Figure). The lowest counts of carotid plaques were observed in individuals with a low parental plaque load, low CAD PRS, low Framingham risk score, or high ideal CVH score in combination with low cumulative neighbourhood disadvantage.

**Figure.**
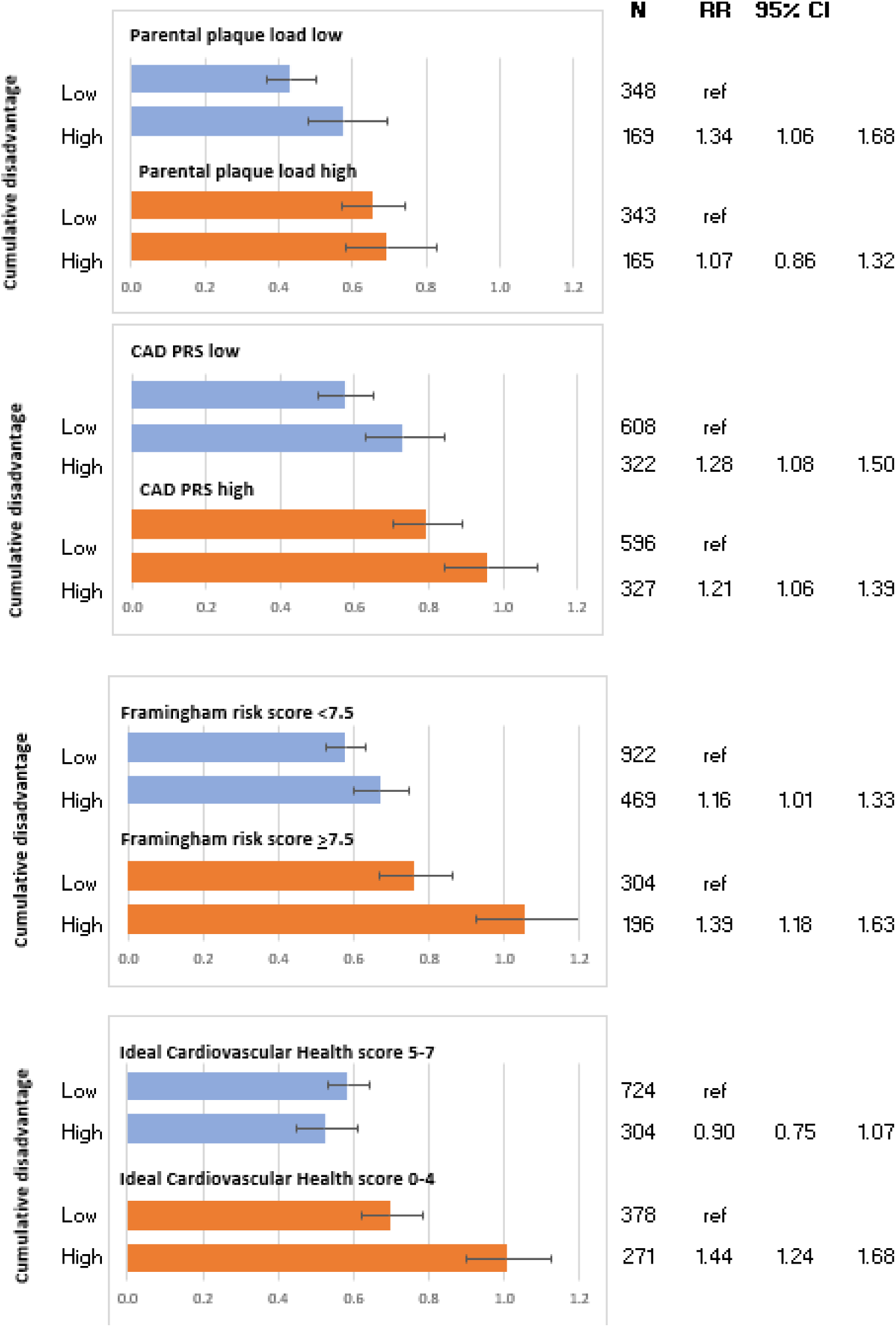
Life course cumulative neighbourhood disadvantage between mean ages 10.7 (SD 5.0, range 3-18) and 48.7 (SD 5.0, range 41-56) years in relation to the national mean (low <0, high >0) and the rate of carotid plaques by the level of cardiovascular risk factors: parental plaque load, CAR PRS (polygenic risk score for coronary artery disease), Framingham risk and Ideal Cardiovascular Health risk scores. The bars denote plaque rates (mean number of carotid plaques in each subgroup) and error bars their 95% confidence intervals. Numeric table on the right panel shows the rate ratios.

In comparison, the carotid plaque count in individuals who had been exposed to high neighbourhood disadvantage was 1.61-fold (95% CI 1.28-2.03) in combination with a high parental plaque load, 1.68-fold (95% CI 1.45-1.95) in combination with a high CAD PRS, 1.82-fold (95% CI 1.56-213) in combination with high Framingham risk score and 1.72-fold (95% CI 1.49-1.98) in combination with low ideal CVH score.

High neighbourhood disadvantage was associated with increased plaque count both in individuals with a low as well as high risk status. The only exception was a lack of association between neighbourhood disadvantage and carotid plaque count among those with ideal CVH score (test of interaction P<0.001). This observation again indicated that the ideal CVH score or its components could play a role in mediating the link between neighbourhood disadvantage and carotid plaque.

To further test this hypothesis, we performed a formal mediation analyses examining the role of each ideal CVH score component separately and in combination as a potential mediator of the association between neighbourhood disadvantage and carotid plaque. The results are shown in Table 4. The column *indirect effect* gives an estimate of the mediation, *i.e.* how large proportion of the association between neighbourhood disadvantage and carotid plaque is mediated by ideal CVH health. Adjusted for age and sex, the ideal CVH score components in combination mediated almost half of the total effect and more than one third after further adjustment for parental SES, own education and marital status. Of the individual components, non-smoking (15%) and ideal blood pressure (6%) were the strongest mediators.

**Table 4.** Total, direct and indirect (through Ideal Cardiovascular Health score components separately and in combination) effect of neighbourhood disadvantage* on the the development of carotid plaques.

| Mediator | N | Total effect |  |  | Direct effect |  |  | Indirect effect |  |  |  |
| --- | --- | --- | --- | --- | --- | --- | --- | --- | --- | --- | --- |
|  |  | RR | lower | upper | RR | lower | upper | RR | lower | upper | % total† |
| Adjusted for age and sex |  |  |  |  |  |  |  |  |  |  |  |
| Ideal blood pressure | 1995 | 1.28 | 1.15 | 1.41 | 1.26 | 1.14 | 1.39 | 1.01 | 1.00 | 1.03 | 5.8 |
| Ideal cholesterol | 1992 | 1.28 | 1.16 | 1.41 | 1.27 | 1.15 | 1.41 | 1.01 | 0.98 | 1.03 | 2.1 |
| Ideal glucose | 1992 | 1.28 | 1.16 | 1.41 | 1.28 | 1.16 | 1.41 | 1.00 | 0.98 | 1.01 | -1.3 |
| Non-smoker | 1900 | 1.26 | 1.14 | 1.40 | 1.22 | 1.11 | 1.34 | 1.04 | 1.01 | 1.07 | 15.2 |
| Physically active | 1882 | 1.24 | 1.12 | 1.37 | 1.22 | 1.11 | 1.35 | 1.01 | 0.99 | 1.04 | 6.2 |
| BMI <25 kg/m2 | 1998 | 1.28 | 1.15 | 1.41 | 1.27 | 1.15 | 1.40 | 1.01 | 0.99 | 1.03 | 3.3 |
| Ideal diet | 1741 | 1.23 | 1.11 | 1.37 | 1.22 | 1.10 | 1.35 | 1.01 | 0.99 | 1.04 | 5.6 |
| Ideal score components in combination | 1677 | 1.23 | 1.10 | 1.37 | 1.12 | 1.03 | 1.23 | 1.09 | 1.02 | 1.17 | 43.1 |
| Adjusted for age, sex, parental SES, education and marital status |  |  |  |  |  |  |  |  |  |  |  |
| Ideal blood pressure | 1903 | 1.21 | 1.09 | 1.35 | 1.20 | 1.08 | 1.33 | 1.01 | 1.00 | 1.03 | 6.5 |
| Ideal cholesterol | 1901 | 1.21 | 1.09 | 1.35 | 1.20 | 1.08 | 1.34 | 1.01 | 0.98 | 1.04 | 5.5 |
| Ideal glucose | 1901 | 1.21 | 1.09 | 1.35 | 1.22 | 1.09 | 1.35 | 1.00 | 0.98 | 1.01 | -1.4 |
| Non-smoker | 1884 | 1.22 | 1.09 | 1.36 | 1.19 | 1.08 | 1.32 | 1.02 | 1.00 | 1.05 | 9.5 |
| Physically active | 1866 | 1.21 | 1.08 | 1.35 | 1.20 | 1.08 | 1.33 | 1.01 | 0.99 | 1.03 | 3.6 |
| BMI <25 kg/m2 | 1905 | 1.21 | 1.09 | 1.35 | 1.20 | 1.08 | 1.34 | 1.01 | 0.99 | 1.03 | 3.8 |
| Ideal diet | 1721 | 1.19 | 1.07 | 1.34 | 1.18 | 1.06 | 1.32 | 1.01 | 0.99 | 1.03 | 4.8 |
| Ideal score components in combination | 1666 | 1.19 | 1.06 | 1.34 | 1.12 | 1.01 | 1.23 | 1.07 | 1.00 | 1.14 | 36.1 |

\*Cumulative disadvantage between mean ages 10.7-48.7 years in relation to the national mean: Low <0, High >0
† Percentages mediated of the total effect by the mediator

## Discussion

Our study demonstrates that cumulative exposure to neighbourhood socioeconomic disadvantage, over 40 years, beginning in childhood, is associated with an increased risk of atherosclerotic carotid artery plaques in mid-adulthood. This association was independent of own or parental socioeconomic position. We additionally found evidence that the association between neighbourhood socioeconomic disadvantage and plaque count was substantially mediated by cardiovascular risk profiles, especially the ideal CVH score and its components. This indicates that behavioural elements (e.g. smoking) may play a key role in explaining the association between cumulative neighbourhood socioeconomic disadvantage and elevated risk of atherosclerotic carotid artery plaque. These observations suggest that long-term exposure to neighbourhood socioeconomic disadvantage is an atherogenic risk factor that operating at least in part through behavioural mechanisms, such as initiation of smoking during childhood/adolescence and/or inability to stop smoking during adulthood. Our findings are in line with the few existing studies that suggest an association of high neighbourhood socioeconomic disadvantage across the life course with cardiovascular disease risk and events in later life^25–28^.

Neighbourhood socioeconomic disadvantage and individuals’ own socioeconomic status have been linked to markers of subclinical atherosclerosis, such as greater carotid IMT and coronary artery calcification (recently reviewed by Hajj et al.^29^). In many previous studies, however, the effect of neighbourhood socioeconomic disadvantage has been attenuated when information on individual socioeconomic status was considered simultaneously, suggesting that the association between neighborhood disadvantage and cardiovascular risk may reflect compositional influences, as opposed to contextual effects of neighborhood environments *per se*. For example, the Atherosclerosis Risk in Communities Study found that lower cumulative neighbourhood socioeconomic status across the life course was associated with greater mean carotid IMT among white participants. However, this association became statistically non-significant when individual-level socioeconomic was considered simultaneously^30^. Similarly, the Cardiovascular Health Study found that while neighbourhood socioeconomic disadvantage was inversely related to prevalent subclinical disease, these associations did not remain statistically significant after adjustment for own socioeconomic position^12^. In contrast, we found that association between cumulative neighbourhood socioeconomic disadvantage and carotid atherosclerosis was only slightly attenuated after adjustment for individual-level socioeconomic position. The stronger independent effects of neighbourhood socioeconomic disadvantage on subclinical atherosclerosis observed in our study are likely attributable to our ability to capture cumulatively life-course exposures across a nearly 40-year prospective follow-up.

Atherosclerosis frequently begins early in life. In line, we have previously demonstrated that childhood smoking and dyslipidaemia predict the development of carotid plaque in adulthood independent of the adult risk status^31^. Furthermore, growing evidence supports an association between early risk factor exposure and increased risk of subsequent cardiovascular events later in life^4,5^. Therefore, the harmful effects of neighbourhood socioeconomic disadvantage likely begin to operate early in life. In a cohort of Montreal children, followed from 1976 to 2010, males from disadvantaged neighbourhoods during childhood were more than 2-times more likely to develop a cardiovascular event compared to males from more advantaged neighbourhoods^32^. These observations indicate that living in socioeconomically disadvantaged areas begin to shape health from childhood onwards. Life-style related behaviours (such as smoking initiation, physical activity, and diet) likely play a major role in explaining these links. We have previously shown in this cohort that high neighbourhood socioeconomic disadvantage is characterised by decreased fruit and vegetable intake as early as age 6 years, as well as decreased physical activity, and increased prevalence of daily smoking from adolescence onwards^27^. Many life-style behaviours are moulded early in the life course, as evidences by significant tracking of physical activity^33^ and dietary patterns^34^ from childhood to adulthood.

The strengths of our study included prospective design with a follow-up of over 38 years from childhood to adulthood and detailed assessment of atherosclerotic carotid artery plaques in adulthood. The possibility to measure neighbourhood socioeconomic disadvantage with objective high-resolution data of all residential locations with dates of moves linked to neighbourhood socioeconomic disadvantage by place and time over the whole follow-up is unique. In other cohorts, neighbourhood disadvantage has been measured either at baseline^3^ or with addresses at different age periods^11,25,26,30,32^. In addition, the area unit in our study, a 250m*250m grid, is likely to capture the variation in the characteristics of the residential location more accurately than large areas, such as counties^1^.

There are some limitations. Since our measurement of neighbourhood socioeconomic disadvantage was limited to three features (education, unemployment, and mean household income), we do not know, what characteristics of neighbourhoods associating with disadvantage could explain the linkage. Potential explanations include, *eg.* access to tobacco retail outlets, traffic noise, exposure to peer influence in schools, lack of usable green space and pollution^14,35–37^. The cohort was racially homogeneous (all white Europeans) and done in a single country. These facts potentially restrict the generalisability of the findings.

In conclusion, these longitudinal data show that exposure to neighbourhood socioeconomic disadvantage beginning in childhood is associated with subclinical atherosclerosis in midlife independently of individual socioeconomic status. Behavioural risk factors seem mediate this link partially. If these observational data reflect causal links, the results highlight the importance of early interventions targeting both social environments and health behaviours to mitigate cardiovascular risk.

## Data Availability

Data are available upon reasonable request

